# Evaluation of spike protein antigens for SARS-CoV-2 serology

**DOI:** 10.1101/2021.01.27.21250382

**Authors:** Suraj Jagtap, K Ratnasri, Priyanka Valloly, Rakhi Sharma, Satyaghosh Maurya, Anushree Gaigore, Chitra Ardhya, Dayananda S. Biligi, Bapu Koundinya Desiraju, Uma Chandra Mouli Natchu, Deepak Kumar Saini, Rahul Roy

## Abstract

**Background:** Spike protein domains are being used in various serology-based assays to detect prior exposure to SARS-CoV-2 virus. However, there has been limited comparison of human antibody titers against various spike protein antigens among COVID-19 infected patients.

**Methods:** We compared four spike proteins (RBD, S1, S2 and a stabilized spike trimer (ST)) representing commonly used antigens for their reactivity to human IgG antibodies using indirect ELISA in serum from COVID-19 patients and pre-2020 samples. ST ELISA was also compared against the EUROIMMUN IgG ELISA test. Further, we estimated time appropriate IgG and IgA seropositivity rates in COVID-19 patients using a panel of sera samples collected longitudinally from the day ofonset of symptoms (DOS).

**Results:** Among the four spike antigens tested, the ST demonstrated the highest sensitivity (86.2%; 95% CI: 77.8-91.7%), while all four antigens showed high specificity to COVID-19 sera (94.7-96.8%). 13.8% (13/94) of the samples did not show seroconversion in any of the four antigen-based assays. In a double-blinded head-to-head comparison, ST based IgG ELISA displayed a better sensitivity (87.5%, 95%CI: 76.4-93.8%) than the EUROIMMUN IgG ELISA (67.9%, 95% CI: 54.8-78.6%). Further, in ST-based assays, we found 48% and 50% seroconversion in the first six days (from DOS) for IgG and IgA antibodies, respectively, which increased to 84% (IgG) and 85% (IgA) for samples collected ≥22 days DOS.

**Conclusions:** Comparison of spike antigens demonstrates that spike trimer protein is a superior option as an ELISA antigen for COVID-19 serology.

**Highlights:**

- Spike trimer displays the highest antibody titer in SARS-CoV-2 infections among spike protein antigens.
- Spike trimer IgG ELISA displays a sensitivity of 50% within six days and 86.2% after 14 days from onset of symptoms.
- IgA and IgG responses to spike trimer antigen were comparable and concomitant in time after infection.
- 16% (IgG) and 15% (IgA) of COVID-19 RT-PCR positive patients did not seroconvert even after 21 days from onset of symptoms.

## Introduction

COVID-19 pandemic caused by the Severe Acute Respiratory Syndrome Coronavirus 2 (SARS-CoV-2) has already crossed 97 million detected cases and over 2 million deaths worldwide till date [1]. As the pandemic continues, we need accurate and sensitive tests to assess the prevalence, disease burden and the level of population immunity against the virus. With the introduction of multiple vaccines and several ongoing vaccine trials, identifying prior exposure or immunogenicity of the vaccine in individuals becomes critical to the development of vaccination and public health strategies.

Nucleic acid-based tests that detect viral RNA are widely used to diagnose active infection in SARS-CoV-2 infected individuals [2,3]. In contrast, immunological tests like serological assays detect the level of human antibody response to the infection in symptomatic as well as the large fraction of asymptomatic infections [4–7]. Immuno-assays detect antigen-specific IgA, IgM and IgG immunoglobulins (antibodies) from body fluids like serum or plasma. Viral antigen-specific antibodies can be detected in SARS-CoV-2 exposed individuals within 5-12 days post-onset of symptoms (POS) for IgM and IgA antibodies and 14 days for IgG antibodies [8,9]. IgG antibodies are long-lived, detectable for at least 8 months, making them promising recent and long term markers of exposure to SARS-CoV-2 [10].

Serological assays with nucleocapsid or spike protein of SARS-CoV-2 as capturing antigen have been widely developed and reported, as these antigens are highly immunogenic [11,12]. Nucleocapsid protein bound to the viral RNA is significantly conserved among the coronaviruses, contributing to false positives in immunoassays [13,14]. The spike (S) protein decorates the exterior of SARS-CoV-2 virus and helps in host cell entry [15–17]. Anti-spike antibodies also demonstrate high virus neutralization efficacy [18,19]. The spike glycoprotein is a clove-shaped trimeric protein with each unit consisting of the S1 head and the S2 stalk. The Receptor Binding Domain (RBD) of the S1 head is responsible for binding to the ACE2 receptor on the cellular membrane, initiating cell entry [16,20]. Due to the large size (153 KDa) of S protein and challenges with achieving proper protein folding, it is difficult to express in bacteria and provides poor yields using traditional mammalian expression systems. Capture antigens used for serological assays should be easy to express and purify, with high yield and stability. Recently, the prefusion state of SARS-CoV-2 spike trimer (ST) protein was stabilized by the addition of 6 prolines that improved thermal stability and expression yield in mammalian cell suspension culture, making it a promising antigen for SARS-CoV-2 antibody assays [21].

In this study, we evaluated the ST protein as a potential capture antigen for ELISA and compared it with different subunits of S protein, namely, S1, S2 and RBD to assess IgG antibody titers in SARS-CoV-2 positive and pre-pandemic sera. We also used ST protein to elucidate IgG and IgA antibody response dynamics with time-stratified samples (≤6, 7-14, 15-21 and ≥22 days POS). Further, we benchmarked the ST protein ELISA against an FDA approved (EUROIMMUN) serology ELISA kit.

## Materials and methods

### Sample collection

For COVID-19 samples, 1-2 ml of blood was drawn from patients who had tested positive for SARS-CoV-2 by RT-PCR test. One set of serum samples (n=69) of COVID-19 patients were obtained from individuals hospitalized at Bangalore Medical College and Research Institute (BMCRI) between April-May 2020. All these samples were collected ≥15 days post-onset of symptoms or RT-PCR positivity (POS/RT). A separate set of serum samples (n=100; collected between March-August 2020) were obtained from COVID-19 biorepository of Translational Health Science and Technology Institute, Delhi (THSTI) that were time stratified along the course of COVID-19 infection (25 samples each from day 0-6, 7-14, 15-21 and ≥22 days POS). For COVID-19 negative controls, serum samples collected during 2018-19 from healthy donors (n=33) and dengue patients (n=61, Panbio dengue IgG/IgM capture ELISA kits, 01PE10/01PE20) and stored in -80°C were used. We also tested the control samples for Influenza A/B antibodies (Immunolab Influenza A/B IgG ELISA, ILE-IFA01/ILE-IFB01). Informed consent from patients was received as per study protocols approved by the Institute Ethics Committees of the institutes where samples were collected and assays were performed.

### Protein expression and purification

The plasmids for RBD (pCAGGS vector containing the human codon-optimized RBD (amino acids 319-541) SARS-CoV-2, Wuhan-Hu-1 spike glycoprotein, GenBank: MN908947; a gift from Florian Krammer, Mount Sinai, New York) and the ST protein (HexaPro; a gift from Jason McLellan, University of Texas, Austin) were purified from DH5α strain of *E. coli*. The plasmids were transfected into Expi293F cells grown using Expi293 expression medium. The cells were grown in a cell culture incubator (37°C, ≥80% relative humidity and 8% CO_2_) on an orbital shaker (130 rpm). Transfection was done at a final cell density of 3×10^6^ viable cells/ml using ExpiFectamine 293 transfection kit as per kit’s instructions. Culture media were harvested after 5 days post-transfection. ST protein was purified using Gravity Flow Strep-Tactin XT resin, whereas RBD protein was purified by HisTrap FF Crude histidine-tagged column on AKTA-Start FPLC system and concentrated using Centricon filter spin columns. Average purified protein yields of 12 mg/l and 61 mg/l were achieved for ST and RBD protein respectively. Spike protein subunits S1 (Native Antigen, REC31806) and S2 (Native Antigen, REC31807) for SARS-CoV-2 were commercially purchased.

### Human IgG and IgA SARS-CoV-2 ELISA

Microtiter plates (Thermo Fisher, 442404) were coated with 50 μl antigen at a concentration of 5 μg/ml in 0.1 M sodium carbonate-bicarbonate buffer (pH 9.6) and incubated overnight at 4°C. Excess unbound antigen was removed by washing the wells thrice with 200 μl wash buffer (0.1% Tween 20 in PBS) using an automated plate washer (Tecan HydroFlex). After washing, 100 μl of blocking buffer (10 mg/ml BSA, 0.05% Tween 20 in PBS) was added to the wells and plates were incubated at room temperature (RT) for 1 h with gentle shaking, followed by washing. Serum samples (50 μl) diluted 1:100 times in PBS with 1 mg/ml BSA were added to the wells. After 30 min of incubation at RT, plates were washed 5 times with 300 μl of wash buffer. 50 μl of horseradish peroxidase-conjugated goat anti-Human IgG (GeNei, HPO2) or IgA specific (Sigma-Aldrich, A0295) antibody diluted 1:3000 in PBS, 0.1 mg/ml BSA, and 0.05% Tween 20 was added to the wells and incubated at RT for 30 min. Excess antibody-enzyme conjugate was removed by washing the wells 5 times with 300 μl wash buffer. 50 μl of chromogenic tetramethylbenzidine (TMB) substrate was added, and plates were incubated in the dark with constant shaking. The reaction was stopped after 10 min by adding 50 μl of stop solution (8.5 M acetic acid and 0.5 M sulfuric acid). Absorbance was measured at 450 nm using a microplate reader (Thermo Scientific Varioskan Flash). Background signal for each sample was estimated by running the same assay without any antigen coating. Corrected OD value was obtained by subtracting the background signal for each sample from its respective OD value in the presence of the antigen. The cut-off value was calculated based on the mean and standard deviation (SD) of the control samples’ OD values as mean + 3SD.

Antigen concentration for ELISA was determined by titrating the antigens at different concentrations till signal saturation. The ST reactivity to SARS specific antibodies was tested with an ELISA titration of the SARS Spike specific antibody CR3022 (Native Antigen MAB12422, Supplementary Figure 1). Diluted serum was titrated, and IgG ELISA was performed using ST with COVID-19 positive and control samples to determine the optimal sera dilution (Supplementary Figure 2). We selected 1:100 sera dilution for performing all the ELISAs based on the high correlation between area under the sera dilution curves and the signal contrast between COVID-19 and control samples (Supplementary Figure 2c-d).

Head-to-head comparison of ST ELISA and EUROIMMUN Anti-SARS-CoV-2 (IgG) ELISA (S1 protein-based serology kit approved by FDA and ICMR, EI 2606-9601 G [22]) was performed in a double-blind format where the experimenters were blind to the RT-PCR and seropositivity results. EUROIMMUN ELISA was performed as per the manufacturer’s instructions.

### Data analysis

All statistical analyses and visualization were done using custom-written python codes and GraphPad Prism software (v8.4.3). Unpaired two-tailed Student’s t-test was used to compare the COVID-19 positive and negative groups. Confidence intervals were calculated using Wilson/Brown’s method [23].

## Results

### Comparison between different spike antigens for human IgG antibodies

We compared four different spike protein antigens (S1, S2, RBD and ST) that represent different protein segments commonly being used to evaluate serum reactivity among SARS-CoV-2 patients (Figure 1a). 94 COVID-19 samples collected ≥15 days DOS/RT, and 94 control samples were tested for the presence of spike-specific IgG antibodies (Figure 1b). We noted that in-house purified RBD and ST performed better than commercially procured S1 and S2 in terms of sensitivity and intensity of the positive sample signal. ST showed the highest sensitivity (86.2%) followed by RBD (69.9%), while S1 and S2 domains showed very low sensitivity (51.5% and 50.0%, respectively) (Table 1). 13 COVID-19 samples were found below the cut-off values for all four antigens.

**Figure 1:**
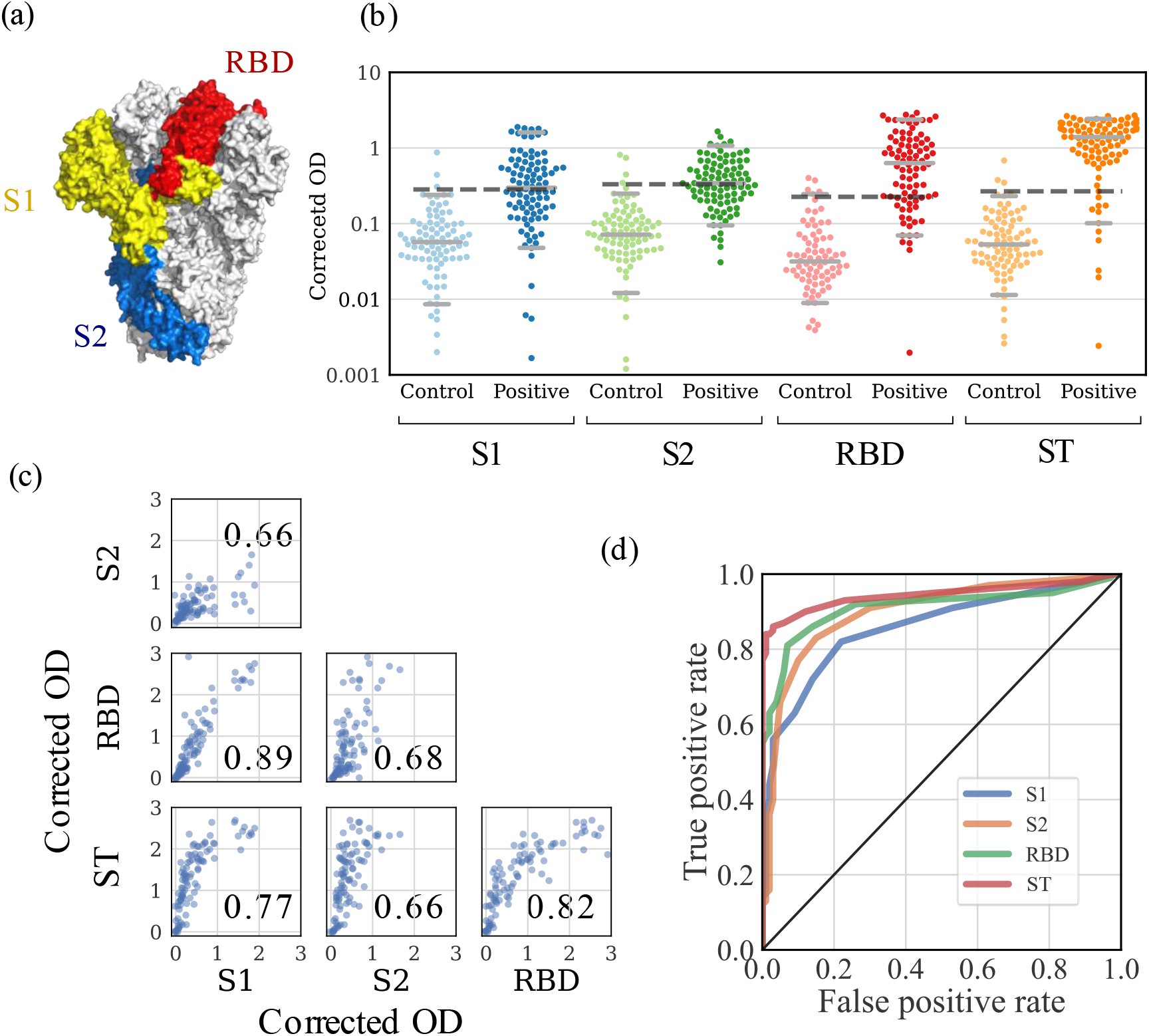
Reactivity of COVID-19 positive (n=94 samples collected ≥15 days from day of -onset of symptoms or RT-PCR positivity) and control sera (n=94) to different antigens (a) Trimeric prefusion spike protein structure (PDB: 5XLR [24]) shows the antigens used in the ELISA (red: RBD, yellow and red: S1 domain, blue: S2 domain, two monomers are represented in white) (b) Corrected OD (450 nm) value for S1, S2, RBD and ST protein for each sample is represented by a point on the scatter plot. Median, 5th percentile and 95th percentile values are shown by horizontal grey lines. Dotted lines indicate the cut-off values. (mean + 3 x standard deviation of corrected OD values of control samples). (c) Correlation between the four antigens. Corrected OD values for each antigen are plotted against the values for the other antigens. Pearson’s correlation coefficient for each pair of antigens is shown on the scatter plots. (d) Receiver operating characteristic curve for each antigen.

**Table 1:**
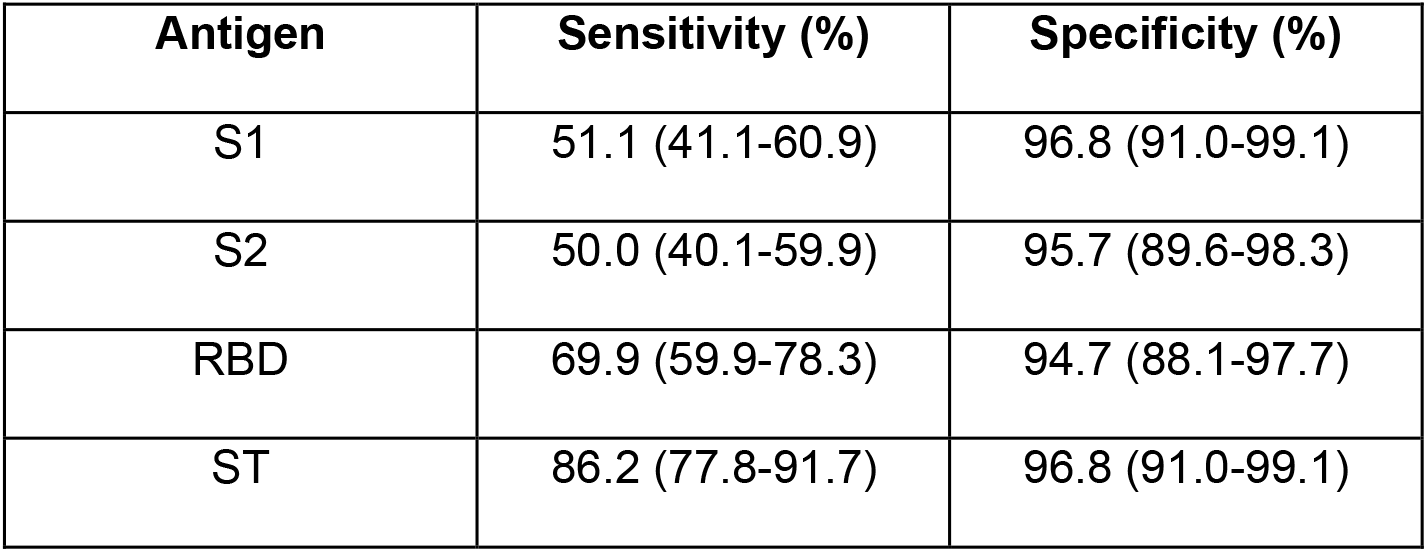
Sensitivity and specificity with 95% confidence intervals (CI) for different spike antigen-based assays.

Among control samples, 64.9% (61/94) were positive for Dengue IgG ELISA, 96.8% (91/94) for Influenza A and 97.8% (92/94) for Influenza B IgG antibodies. This confirmed that these control samples had other detectable virus specific antibodies. Despite this, we obtained specificity >94.7% for all the four antigens (Table 1), suggesting low cross-reactivity of spike antigens s to antibodies for these viruses.

S1, RBD and ST antigens displayed a high correlation among themselves (Pearson’s correlation coefficient (PCC) >0.75 for all cases). S2 subunit displayed a lower correlation (PCC 0.66-0.68) to others. This is likely because the structurally buried S2 domain in the spike protein is less accessible to the antibodies. Nevertheless, the significant correlation across all four antigens (Figure 1c, PCC >0.65), suggests that antibody responses across all spike antigens are consistent and that they are all detectable in our ELISA platform.

ST detected higher OD values, possibly because of the larger number of epitopes available on the full-length soluble trimeric protein. It also displayed the highest contrast between COVID-19 positive and control groups (Figure 1b). Receiver operating characteristic curves indicates that ST is the best candidate for serological testing with the highest area under the curve (Figure 1d). Further, when the ST-based ELISA was repeated in 93 samples (56 COVID-19 positive samples and 37 control samples) in a blind manner, we obtained highly reproducible values (PCC=0.986, Supplementary Figure 3).

### Comparison with commercial IgG serology kit

Corrected OD values obtained from ST-based ELISA and ratios obtained from EUROIMMUN test show a good correlation in a double-blind comparison (Figure 2). However, the ST-based ELISA (87.5% (95% CI: 76.4-93.8%)) performed better than the EUROIMMUN IgG kit (67.9% (95% CI: 54.8-78.6%)) in terms of sensitivity (for samples ≥15 days DOS/RT). Our results are comparable to values reported by the internal validation report of EUROIMMUN (61.1% for 11-21 days DOS and 81.1% for >11 days post PCR positivity [22]) and lesser than those obtained by others (84.4% [25], 89.5% [26]). Specificity values of both methods were comparable - 94.6% (95% CI: 82.3-99.0%) for ST-based ELISA and 97.3% (95% CI: 86.2-99.9%) for EUROIMMUN kit.

**Figure 2:**
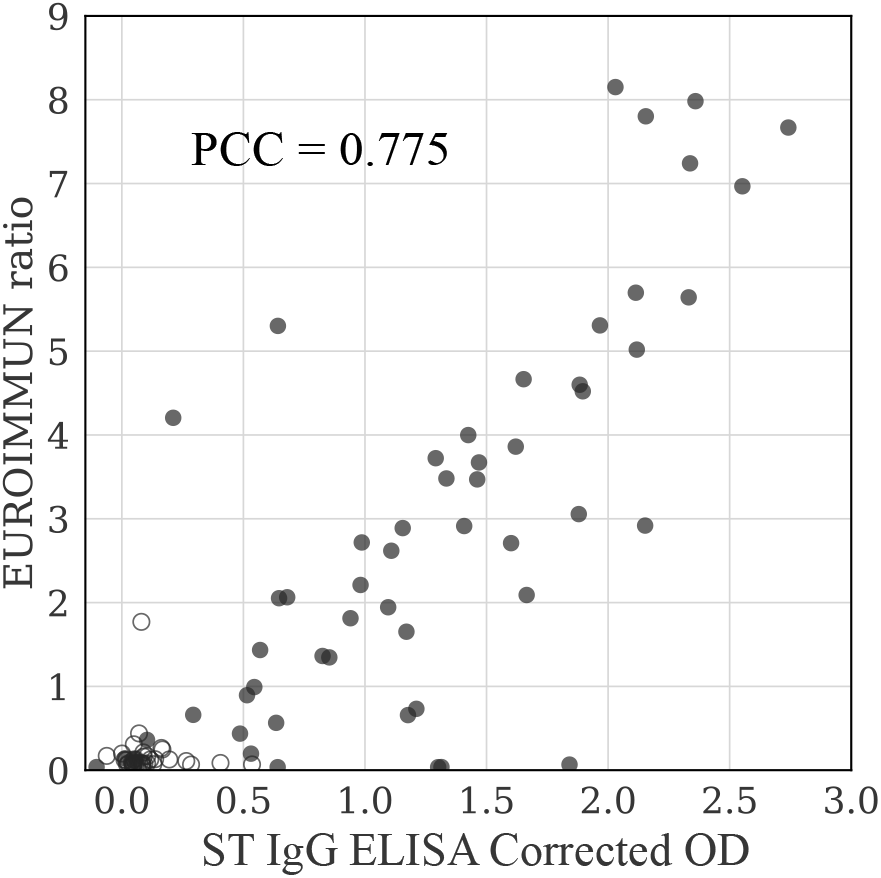
Head-to-head double-blinded comparison of EUROIMMUN Anti-SARS-CoV-2 ELISA (IgG) kit and ST ELISA. COVID-19 positive samples (filled circles) and control samples (empty circles) are plotted and Pearson’s correlation coefficient (PCC) between the two test values is indicated for the COVID-19 positive samples.

### Dynamics of IgG and IgA response in SARS-CoV-2 infections

We performed ELISA assays for spike specific IgG and IgA antibodies in sera collected at different time points (≤6, 7-14, 15-21 and ≥22 days POS) using ST protein. Median OD values of IgG and IgA response increased with time (Figure 3a-b; IgG: 0.21, 0.69, 1.40, 1.57, IgA: 0.26, 0.49, 0.77, 1.06 for the respective time intervals). Also, the proportion of patients who demonstrated seroconversion increased with time (Figure 3c; IgG: 48.0%, 64.0%, 84.0%, 84.0%, IgA: 50.0, 69.6, 72.7, 85.0% for the respective time intervals). In both cases, about 50% of the patients had seroconverted within 6 days from DOS. These findings are in line with previous reports that have shown median time for seroconversion based on spike specific IgG antibodies to be 14 days POS [9,27,28]. A high correlation (PCC = 0.76, Figure 3d) was observed between ranks of IgG and IgA values for each sample suggesting a comparable and concomitant response to the infection.

**Figure 3:**
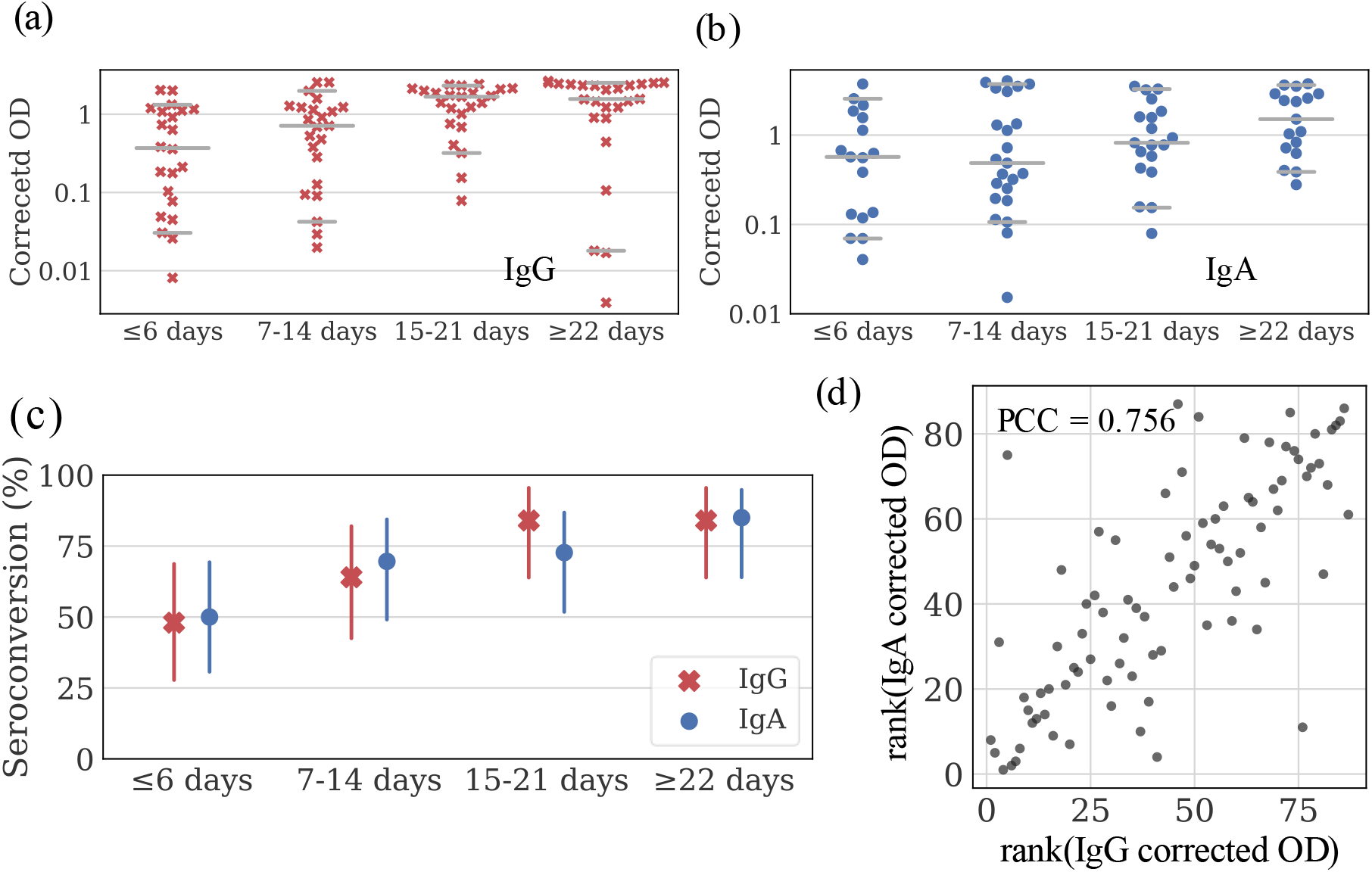
Dynamics of ST protein-specific IgG and IgA response. (a-b) Corrected OD values of COVID-19 positive samples collected at different time points post-onset of symptoms (n=25 for each time interval). Value for each sample is represented by a point on the scatter plot. Median, 5th percentile and 95th percentile values are shown by horizontal gray lines. (c) Percentage seroconversion of patients based on IgG and IgA response to ST protein ELISA as a function of time. Vertical lines show 95% confidence intervals (d) Comparison between ranks of the IgG and IgA corrected OD values.

Some individuals did not show detectable spike-specific antibodies (16% for IgG, 15% for IgA) even after 21 days from DOS. This could be due to asymptomatic/mild infection which has been reported to display low antibody titers [6]. We cannot rule-out false-positive PCR results contributing to some of these cases.

## Discussion

The trimeric spike protein from SARS-CoV-2 is critical for cellular entry and is prominently displayed on the virus. Our results revealed that ST protein displayed better reactivity to COVID-19 positive sera when compared to S1, S2 and RBD spike subunit proteins consistent with the larger number of accessible antibody epitopes on ST. While we did not perform neutralization assays, previous studies have shown that the antibodies against RBD are strongly correlated with neutralization of the virus [18,19,29]. Several other regions of the Spike protein, including a region away from receptor binding site [30], S1 domain [31] and S2 domain [32–34] are targets of neutralizing antibodies. Therefore, a high ELISA signal against the ST protein is suggestive of higher levels of a broader spectrum of virus-neutralizing antibodies.

IgG and IgA antibody dynamics show seroconversion in about half of the patients within 6 days POS (also reported earlier, [9,27,35]). Considering the low sensitivity of many existing rapid antigen tests [36–38], a combination of the rapid antigen detection with antibody tests can be employed to increase the detection efficiency of COVID-19 cases during early infection.

Some COVID-19 RT-PCR positive samples did not show reactivity against any of the four antigens. Several other studies have reported limited seroconversion at the time of sera collection [6,39,40]. Nevertheless, highly sensitive serology assays can be critical in determining the sero-prevalence in a community as well as help in assessing the immune response/ immunogenicity of vaccines including the temporal course of antibody responses. Our results show that the human antibody response is consistent among the spike antigens, and the spike trimer maybe the best choice of antigen for COVID-19 serology.

## Data Availability

The authors confirm that the data supporting the findings of this study are available within the article and its supplementary materials. Additional raw and derived data supporting the findings of this study are available from the corresponding author [RR] on request.

## Author contributions

SJ, RK, PV and RR conceptualized the study; CA, DSB, SJ, RK, PV and RR devised methods, collected samples and clinical data; SJ, RK, PV, RS, SM, AG, CA, DKS, DSB and RR provided reagents and analysed samples; SJ, RK, PV, RR and BKD curated and/or analysed data; SJ, RK, PV, UCMN and RR drafted the manuscript with inputs from others; RR coordinated and supervised the study. All authors approved the final draft of the manuscript.

## Acknowledgements

This research has been conducted with the contribution of NCR Biotech Science Cluster BIOREPOSITORY, DBT India consortium for COVID-19 Research and RGUHS-IISc Dengue biomarker research consortium. We thank Florian Krammer, Jason McLellan, Raghavan Varadarajan, Sameer Malladi for providing useful reagents; Sunaina Banerjee and Rohit Dutta for valuable discussions and Khantesh Agrawal for anonymizing the samples. This work was supported by the Indian Institute of Science and CSR funding by Capgemini India. Funding agencies had no role in the design, execution, or interpretation of the study.

