## Supplementary material for "Evaluation of spike protein antigens for SARS-CoV-2 serology"

*Jagtap et al. 2021*

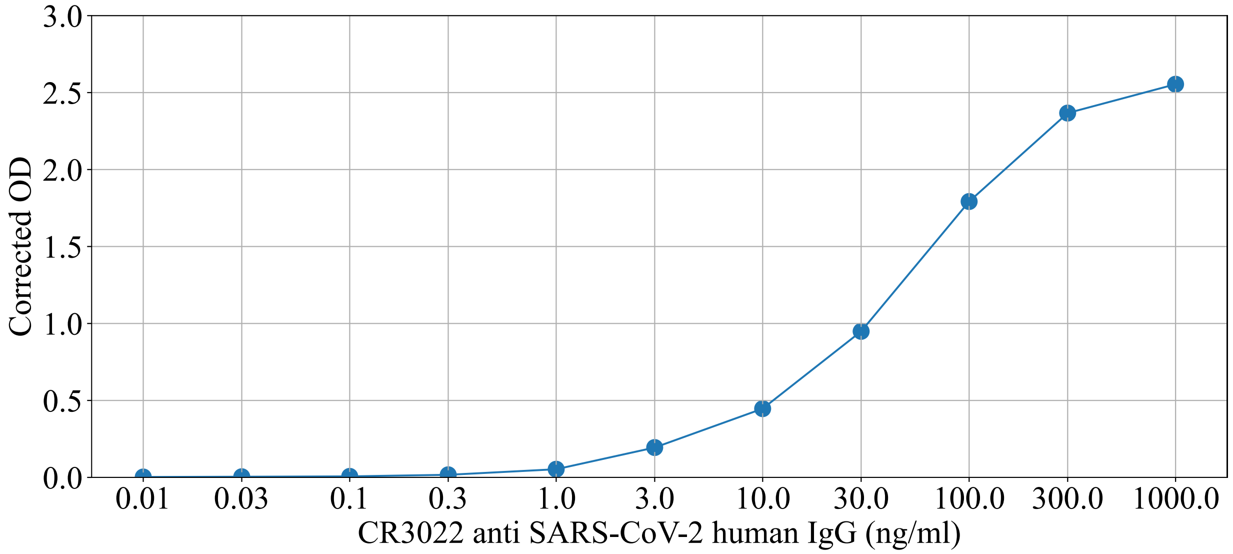


Supplementary Figure 1: Spike protein-specific antibody CR3022 binding to ST protein.

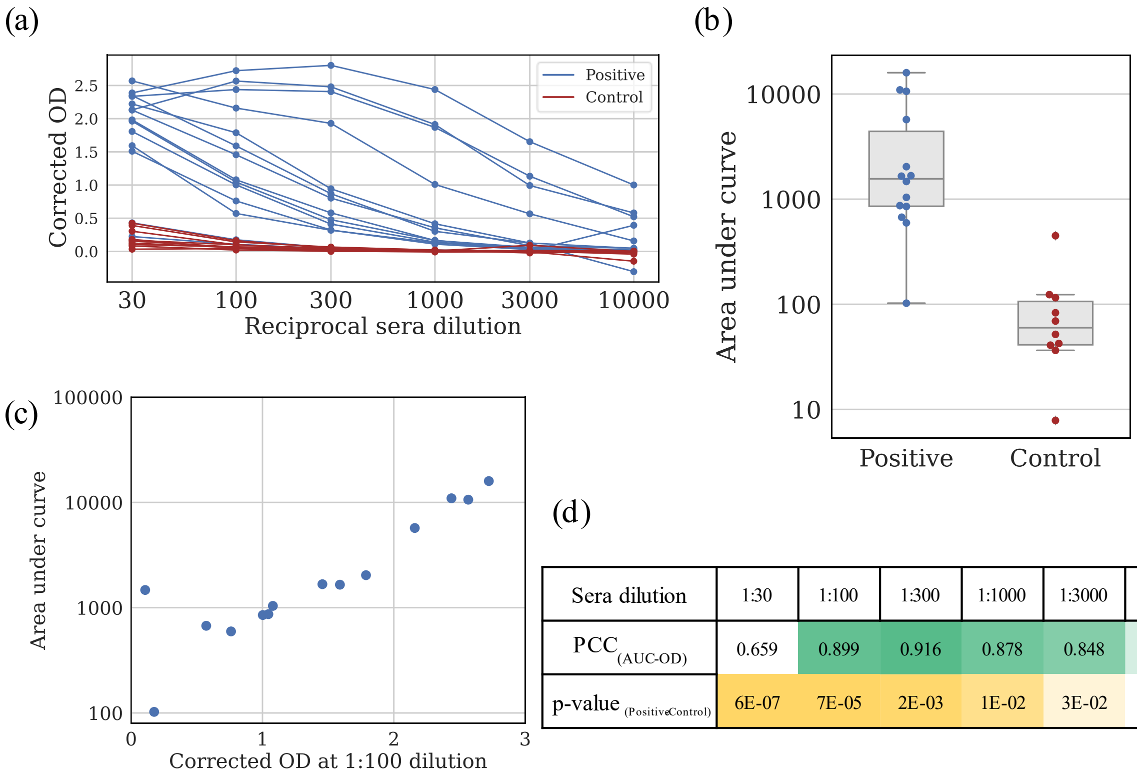


| Sera dilution | 1:30 | 1:100 | 1:300 | 1:1000 | 1:3000 | 1:10000 |
|---|---|---|---|---|---|---|
| $PCC_{(AUC\text{-}OD)}$ | 0.659 | 0.899 | 0.916 | 0.878 | 0.848 | 0.724 |
| p-value $_{(Positive\text{-}Control)}$ | 6E-07 | 7E-05 | 2E-03 | 1E-02 | 3E-02 | 4E-02 |

Supplementary Figure 2: Optimization of sera dilution. (a) Sera reactivity at different sera dilutions. The x-axis indicates reciprocal dilutions of the sera. Corrected OD values at 450 nm for COVID-19 positive samples are shown in blue and control samples in red (b) Comparison of area under the curve (AUC) for both the groups (c) Correlation between area under the curve and corrected OD measurements at 1:100 sera dilution (d) Row 1 of the table shows Pearson's correlation coefficient (PCC) between AUC and OD measurements at different dilutions. Row 2 indicates p-value of the unpaired two-tailed Student’s t-test between positive and control sera at different sera dilutions. Darker shades of green represent higher PCC values, and darker yellow shades indicate more significant p-values.

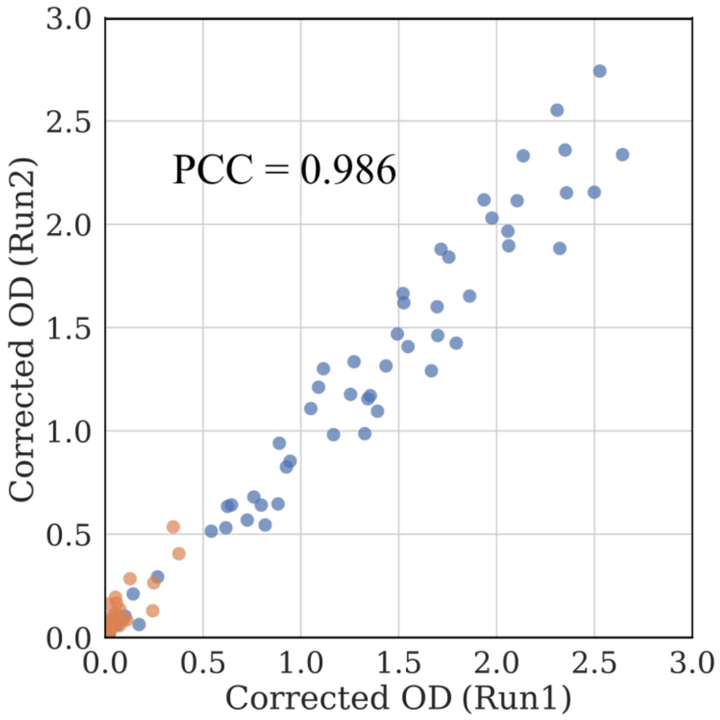


Supplementary Figure 3: Data reproducibility. Corrected OD values from two independent ST-based ELISA runs are reported (PCC = 0.986). COVID-19 positive samples (n=56) are shown in blue and control samples (n=37) are shown in orange
